# Effects of Tranexamic Acid in Reducing Blood Loss in Patients Undergoing Coronary Artery Bypass Surgery: A Systematic Review

**DOI:** 10.1101/2020.07.26.20162388

**Authors:** Ahish Chitneni, Suhani Dalal

## Abstract

Tranexamic acid is a medication typically used to prevent excessive blood loss in the setting of trauma, surgery, and menstrual cycles. Administration of tranexamic acid within the first three hours of trauma has been shown to reduce blood loss and mortality in patients. [1] Tranexamic acid is also commonly used during heavy menstrual bleeding or postpartum hemorrhage cases in which administration has been shown to reduce bleeding in patients. [2] Although there have been studies conducted on the effects of tranexamic acid on blood loss during various surgeries, this systematic review aims to understand the effects specifically on patients undergoing coronary artery bypass grafting (CABG), a common procedure known to have significant blood loss. A systematic review using the PubMed database on the use of tranexamic acid in blood loss for patients undergoing CABG was conducted with the hypothesis that tranexamic acid administration reduces perioperative and postoperative blood loss. This systematic review includes 8 papers from the years 1997-2018 which report on a total of 712 patients. Although significant large, randomized, control studies on the topic are still pending, this systematic review of studies conducted thus far can conclude that tranexamic acid has a positive effect on reducing blood loss in patients undergoing CABG.

## Introduction & Background

Coronary artery bypass surgery (CABG) is a surgical procedure conducted to improve blood flow to the heart. Typically, the surgery is conducted when major coronary arteries supplying the heart are blocked or narrowed and limit functioning of the heart. CABG can be done with the use of a cardiopulmonary bypass machine (on-pump) or done without (off-pump). [3] CABG is utilized in patients with obstructive coronary disease or in emergent situations such as patients with myocardial infarctions to improve blood flow to the heart and provide therapeutic relief. [4]

Despite advances in surgical techniques and procedures, bleeding during cardiac surgery is a fairly common occurrence. Perioperative bleeding during surgery has been shown to significantly correlate with morbidity and mortality in patients. In fact, patients that undergo reoperation for bleeding after cardiac surgery have been shown to have a 4.5-fold higher overall mortality than patients who do not. [5] Given the fact that tools that accurately predict blood loss during surgery do not exist, it is important to limit the amount of blood loss that occurs during the surgical procedure. [6]

Tranexamic acid (trans-4-(aminomethyl) cyclohexanecarboxylic acid) is a derivative of lysine that is an antifibrinolytic that works by reversibly blocking the lysine binding sites on plasminogen molecules. In addition, it works as an inhibitor of tissue plasminogen activator (TPA). Both of these mechanisms work to inhibit the activation of fibrinogen and thus has an anti-fibrinolytic effect. [7] Recently, there has been widespread interest in the role of tranexamic acid in reducing blood loss in patients undergoing surgery. The CRASH-2 study was the largest trial conducted studying the effects of tranexamic acid in reducing blood loss for trauma patients. The study was a randomized, placebo-controlled trial that showed that mortality and the risk of bleeding was significantly reduced in trauma patients who received tranexamic acid compared to the placebo. [8]

Given the clear positive effects of tranexamic acid seen in studies for trauma patients, this review seeks to understand the role it plays in CABG, one of the most common cardiac surgeries performed in the United States. An up-to-date systematic review was conducted on all of the studies in the PubMed database that observed the effects of tranexamic acid in reducing blood loss perioperatively and postoperatively in patients undergoing the procedure. It is hypothesized that tranexamic acid will have similar effects on this patient population as it did with trauma patients undergoing rescue surgery.

## Review

### Methods

The PubMed database was searched using the keywords “tranexamic acid AND CABG” and the keywords “tranexamic acid AND blood loss AND CABG”. No timeframe limitations were placed on the search. Any randomized, placebo-controlled clinical trial that studied the effects of tranexamic acid on perioperative and postoperative blood loss in patients undergoing CABG were included in the review. Both on-pump and off-pump CABG were included in this systematic review.

Any studies that were also systematic reviews, meta analyses, or poster presentation cases were not included in the final list of studies that underwent review. All articles that included the study of other antiplatelet or antifibrinolytic agents alongside tranexamic acid were excluded from the review. Articles that studied other procedures or CABG and other procedures were excluded. Publications that were unavailable in English were excluded.

From the initial search results, 58 unique studies were identified as being of relevance to this review. After further analysis of each individual study, 8 unique studies were included in this systematic review. This systematic review includes 8 papers from years 1997-2018 which report on a total of 712 patients.

### Results

From 58 unique potential studies found in the PubMed literature search, 8 articles fulfilled the eligibility criteria for this systematic review. The studies consisted of randomized, placebo-controlled trials of the use of tranexamic acid versus placebo in patients undergoing CABG. The use of tranexamic acid to control bleeding in patients with trauma, postpartum hemorrhage, and heavy menstrual bleeding has been investigated in depth and found to play a significant role in the reduction of blood loss. [2,8] In more recent clinical practice, evidence suggests the use of tranexamic acid in cardiac surgeries can reduce blood loss and thus reduce postoperative transfusions and complications. [9]

Of the 8 studies reviewed, all showed a decrease in blood loss in patients undergoing CABG when compared to a placebo. 6 of these studies showed a statistically significant decrease in perioperative and postoperative blood loss with the administration of tranexamic acid when compared to a placebo. The included studies consist on 7 that review on-pump CABG, and one that studies tranexamic use in off-pump CABG. The studies quantified volume of blood loss, perioperative and postoperative transfusion requirement, hemoglobin, hematocrit, and D-dimer levels to assess the effectiveness of antifibrinolytics in decreasing blood loss after CABG.

#### Chest Tube Drainage

Many studies have shown the advantages of tranexamic acid in decreasing blood loss in trauma patients and postpartum hemorrhages. Several studies have also been conducted to evaluate the effectiveness of tranexamic acid in patients undergoing cardiac procedures such as CABG, which are known to have significant blood loss that can contribute to morbidity and mortality.

Zhang et al. conducted a prospective, randomized, placebo-controlled trial with administration of tranexamic acid compared to placebo in patients undergoing on-pump CABG, with regular follow-up for seven years postoperatively. In the postoperative period, tranexamic acid administration significantly reduced blood loss, measured via chest tube drainage. [10] The 7-year follow-up found no significant difference between the placebo and tranexamic acid group in terms of morbidity and mortality, suggesting that postoperative tranexamic acid administration also carries low risk in the long term.

[10] Similarly, Fawzy et al. found that tranexamic acid administration resulted in a statistically significant decrease in chest tube drainage in the first 24 hours after on-pump CABG, representing a 37% decrease in blood loss. [11] There was a 32% reduction in total blood loss in the postoperative period. [11] Nejad et al. also reported similar findings, citing significantly decreased chest tube drainage in patients treated with tranexamic acid when compared to a placebo following on-pump CABG. [12]

Andreasen et al. studied the use of tranexamic acid in on-pump CABG and found that, although there was less blood loss at 6 hours and 12 hours following surgery in patients that received tranexamic acid, this difference was not statistically significant compared to the control group. [13] The study acknowledges that the small sample size (N = 44, 21 in treatment group and 23 in control group) may contribute to the lack of statistical significance. [13] However, this may also indicate that there is greater importance in achieving surgical hemostasis rather than tranexamic acid administration for reducing overall blood loss in patients. [13] Mirmohammadsadeghi et al. also found decreased average blood loss in patients who underwent treatment with tranexamic acid, but the decrease was not statistically significant. [14]

Hosseini et al. studied the use of tranexamic acid in off-pump CABG of 71 patients, a relatively newer procedure. The study showed that there is also significantly decreased blood loss in patients with tranexamic acid versus placebo. [15] However, the study did not find any difference between the two groups when evaluating volume of blood transfusions, hemoglobin, hematocrit, platelets, prothrombin time, and partial thromboplastin time in the postoperative period. [15]

6 studies included in this review, which represented 543 patients, found statistically significant decrease in blood loss after tranexamic acid administration.

#### Allogeneic Red Blood Cell (RBC) Transfusion Requirements

Review of studies has shown that tranexamic acid use can decrease the need for allogeneic RBC transfusions. RBC transfusions are generally indicated in patients with hemoglobin less than 7 g/dl, which can occur after significant blood loss following trauma, hemorrhage, or surgery. One study, Zhang et al., found that the patients who received tranexamic acid required significantly less allogeneic RBC transfusions compared to the placebo group following on-pump CABG. [10] However, all other studies of on-pump CABG in this review indicate that there is no significant difference between the placebo and treatment groups when evaluating transfusion requirements postoperatively.

One study in this review evaluated transfusion requirements in off-pump CABG. It was found that there was significantly decreased allogeneic RBC transfusion required in patients who received tranexamic acid compared to placebo. [15]

#### Hemoglobin, Hematocrit

Hemoglobin and hematocrit are important indicators of hemostatic change that can help determine the need for transfusions. One study showed statistically significant differences in hemoglobin and hematocrit in patients that received tranexamic acid compared to the placebo group. Yanartas et al. found that patients who received tranexamic acid had a statistically significant higher hemoglobin and hematocrit at 12 hours and 24 hours post-operatively. [16] The study found that tranexamic acid plays an important role in decreasing chest tube drainage following on-pump CABG. [16] Other studies in this review found no statistically significant correlation between the hemoglobin and hematocrit and tranexamic acid administration.

#### D-dimer Levels

Pinosky et al. studied the efficacy of antifibrinolytics in reducing blood loss in patients undergoing on-pump CABG, comparing tranexamic acid to epsilon-aminocaproic acid and a placebo. [17] Epsilon-aminocaproic acid (EACA) is an antifibrinolytic that is also a lysine analogue with similar mechanism of action to tranexamic acid. [18] The study found that tranexamic acid significantly reduces blood loss when compared to epsilon-aminocaproic acid and placebo in the first 24 hours postoperatively. [17] The concentration of the D-dimer, a protein fragment resulting from fibrinolysis, was significantly reduced in patients who received tranexamic acid, suggesting that tranexamic acid administration effectively reduces fibrinolysis during and after CABG. This likely contributes to decreased blood loss after CABG.

## Discussion

Tranexamic acid has been widely studied for its use in trauma patients, postpartum hemorrhage, and gastrointestinal hemorrhage to minimize blood loss and stabilize patients. It has been found to reduce mortality due to traumatic bleeding by one-third and is also known to reduce blood loss and transfusion requirements. [5] The positive effects of tranexamic acid in reducing blood loss with minimal side effects makes it the preferred method of bleeding control when compared to other medications. [19] Because it is relatively safe, tranexamic acid can be administered in a wide spectrum of trauma patients to control bleeding. [20]

This systematic review discussed the effects of tranexamic acid administration in patients undergoing CABG. Various parameters such as chest tube drainage, allogeneic red blood cell transfusions, hemoglobin and hematocrit, and D-dimer levels were quantified in several studies to understand the effects of tranexamic acid administration on overall blood loss. Despite the limited number of studies available on this topic, there is strong evidence supporting the positive effects of tranexamic acid in reducing blood loss in patients who undergo CABG. A meta-analysis published by Zhang et al. in 2019 also found decreased blood loss in patients undergoing CABG with the administration of tranexamic acid. [21] The study reported decreased need for allogeneic blood products without increased risk of prothrombotic events. [21] However, the study noted that tranexamic acid administration may contribute to post-operative seizures. [21]

Various parameters were studied in individual studies, but there is overwhelming correlation between tranexamic acid administration and reduced chest tube drainage post-operatively. Volume of blood loss was quantified by measuring chest tube drainage in patients who underwent CABG. The studies analyzed in this systematic review have found that administration of tranexamic acid reduces blood loss post-operatively, and six of the eight studies found that this difference was statistically significant.

Studies have also found that there is decreased need for red blood cell transfusions, higher hemoglobin and hematocrit levels, and decreased D-dimer levels for patients who receive tranexamic acid. Although there is less evidence that there is a statistically significant difference between groups in these parameters, further research can be conducted to increase the sample size and further our understanding of the effects of tranexamic acid.

It is important to recognize that there are limitations in this systematic review. There is the potential for bias when interpreting literature and collecting data for the review. Furthermore, this study only included literature that was available in English, which limits the research that was available. Another limitation to this systematic review is the limited literature available that met the inclusion criteria for this review. It is important to note that this review included data from both on-pump and off-pump CABG procedures. Off-pump CABG is a fairly newer procedure, and thus, there is less available literature on the effects of tranexamic acid in off-pump CABG.

## Conclusions

The publications analyzed in this review show a positive correlation between tranexamic acid and decreased blood loss in patients undergoing CABG. Evidence in several randomized, placebo-controlled clinical trials shows that tranexamic acid can reduce blood loss with minimal side effects and can be beneficial in patients following CABG. The reduction in blood loss due to tranexamic acid may play an important role in decreasing transfusion requirements and post-operative complications. Given that much of this research has been conducted on on-pump CABG procedures, further work can be done to understand the effects of tranexamic in off-pump CABG.

## Data Availability

All data referred to in the manuscript is available

